# Epidemiological Analysis of Common Infections in Rural Pakistan: Implications for Public Health Interventions

**DOI:** 10.1101/2025.02.18.25322363

**Authors:** Shahzad Mahmood, Nauman Ali Chaudary, Jafar Riaz Kataria, Muhammad Naveed Tahir, Maham Noor

**Affiliations:** Allama Iqbal Medical College Lahore, Pakistan; University of the Punjab Lahore, Pakistan; Department of Public Health University of the Punjab Lahore, Pakistan; Lahore Medical and Dental College

## Abstract

Infectious diseases remain a major public health challenge in rural Pakistan due to limited healthcare access and infrastructure. This cross-sectional study examines the prevalence of gastrointestinal, respiratory, and dermatological infections across different age and gender groups in a rural population. Data were collected over 45 days from a basic health unit serving seven villages with a population of 50,000. Patients were categorized based on infection type, gender, and age group. The findings revealed distinct epidemiological patterns: respiratory and dermatological infections peaked in the 15-49 age group before declining in older adults, while gastrointestinal infections were most prevalent in children aged 1-4 years. Males exhibited a slightly higher prevalence of dermatological infections, whereas gastrointestinal and respiratory infections showed minor gender-based variations. The study highlights key risk factors contributing to disease prevalence and underscores the need for targeted public health interventions, improved healthcare access, and preventive measures tailored to specific demographic groups. Future research should incorporate longitudinal data to establish causal relationships and enhance disease mitigation strategies.

**Short Description:** This study focuses on the distribution of infectious diseases including: respiratory tract infections, gastrointestinal tract infections and dermatological infections. It not only explores the disease’s frequency among different genders and among different age groups, but also provides a thematic analysis of policy recommendations to control the diseases in rural Pakistan

## Background

Infectious diseases continue to pose a significant threat to public health, especially in rural areas where healthcare access is limited. Grasping the extent of infectious diseases in rural regions is essential for creating targeted strategies and enhancing healthcare provision. While extensive research has been conducted on infectious disease burdens in urban environments, data on their prevalence in rural communities remains scarce. There is insufficient comprehensive investigation into the spread and effects of infectious diseases within Pakistan’s rural landscape. This research seeks to address the knowledge gap by offering detailed analysis of gastrointestinal, respiratory, and dermatological infection prevalence in rural Pakistan. What is the occurrence rate of gastrointestinal, respiratory, and dermatological infections in Pakistan’s rural areas, and how does it differ across age groups and genders? This study aims to measure the burden of infectious diseases in rural Pakistan and examine the data through the lens of age and gender demographics. We anticipate that the prevalence of infectious diseases in rural Pakistan will show significant variations among different age groups and between males and females. (1) Instead of multiple sites, a basic health unit was chosen which was the sole reporting site for all the patients of 7 villages and covered a population of 50,000 people. The samples were taken for 45 days from the general OPD of the basic health unit and people were segregated on the basis of whether they had gastrointestinal tract, respiratory tract, or dermatological tract infections. Furthermore, this segregation was further divided into male and female categories and further redivided into age-based categories like the number of males with gastrointestinal tract infection in the age brackets of 1 to 9 years and so on. After quantifying and plotting these segregated variables, the risk factors were studied in the discussion and policy recommendations were given for a targeted approach after analyzing the age and gender distribution and epidemiology of different diseases.

A single basic health unit was selected as the sole reporting site for patients from seven villages, covering a population of 50,000 individuals. Data collection was conducted over a 45-day period from the general outpatient department of the basic health unit. Patients were categorized based on the presence of gastrointestinal, respiratory, or dermatological infections. Further stratification was performed according to gender and age groups, such as the number of males with gastrointestinal tract infections in the 1-9 year age bracket. Following quantification and graphical representation of these stratified variables, risk factors were analyzed in the discussion section, and policy recommendations were proposed for a targeted approach based on the age and gender distribution and epidemiology of various diseases.

### Literature Review: Infectious Disease Prevalence in Rural Pakistan

Infectious diseases remain a significant public health concern, particularly in rural areas with limited healthcare access. (2) While extensive research has been conducted on infectious disease burdens in urban environments, data on their prevalence in rural communities is limited. This literature review aims to address this knowledge gap by examining the occurrence rates of gastrointestinal (3), respiratory (4), and dermatological infections (5) in Pakistan’s rural areas, with a focus on age and gender demographics.

Understanding the extent of infectious diseases in rural regions is crucial for developing targeted strategies and improving healthcare provision. Current research has primarily focused on urban settings, leaving a notable gap in our understanding of infectious disease prevalence in rural Pakistan. This study seeks to address this gap by providing a comprehensive analysis of gastrointestinal, respiratory, and dermatological infection prevalence in rural Pakistan.

A cross-sectional study was conducted in a rural area of Pakistan, utilizing a single basic health unit as the sole reporting site for patients from seven villages, encompassing a population of 50,000 individuals. Data collection was carried out over a 45-day period from the general outpatient department. Patients were categorized based on the presence of gastrointestinal, respiratory, or dermatological infections (6), with further stratification according to gender and age groups. The methodology employed a stratified random sampling method to ensure adequate representation of demographic factors. Data collection involved standardized questionnaires, and clinical examinations by trained healthcare professionals. (7) Ethical considerations were addressed through obtaining approval from the institutional review boards and securing informed consent from participants.

Analysis of the collected data revealed distinct patterns across age and gender for different types of infections. Respiratory and dermatological infections demonstrated a peak in the 15-49 age group (8), potentially due to lifestyle factors or changes in immune function, before declining in older adults. Males exhibited a slightly higher prevalence of dermatological infections (9) across most age groups. In contrast, gastrointestinal infections demonstrated a unique pattern, with the highest number of cases concentrated in the 1-4 year age group.(10) This suggests that young children are particularly susceptible to these infections, possibly due to immature immune systems or exposure to pathogens in daycare or preschool settings. A consistent decrease in gastrointestinal infection cases was observed with increasing age after the early childhood peak.(11)

The study’s findings highlight the varying epidemiological factors influencing different types of infections in rural Pakistan.(12) The observed patterns suggest potential influences of lifestyle, occupational exposure, and age-related immune function changes on infection prevalence. The higher prevalence of gastrointestinal infections in young children aligns with the understanding of developing immune systems and potential environmental exposures.

While this cross-sectional study provides valuable insights into the prevalence and distribution of infections in rural Pakistan, it is important to acknowledge its limitations. The study design offers a snapshot of disease burden but limits the ability to establish causal relationships. Additionally, relying on self-reported symptoms introduces the possibility of recall bias.

In conclusion, this literature review underscores the importance of understanding infectious disease prevalence in rural Pakistan. The findings provide a foundation for developing targeted public health interventions and improving healthcare provision in rural areas. Future research incorporating longitudinal data and population-based rates would further enhance our understanding of these infectious disease patterns in rural settings

## Methodology

### Study Design

A cross-sectional study was conducted to assess the prevalence of gastrointestinal, respiratory, and dermatological infections in rural Pakistan. This design allowed for the collection of data at a specific point in time, providing a snapshot of the current disease burden.

### Study Setting

The study was carried out in a selected rural area of Pakistan. A single basic health unit was chosen to ensure a representative sample of the rural population.

### Participant Selection

A stratified random sampling method was employed to select participants. The population was stratified by age groups and gender to ensure adequate representation of these demographic factors. Inclusion criteria encompassed residents of the selected rural area, while exclusion criteria were determined based on specific study requirements.

### Sample Size Calculation

The sample size was calculated using standard statistical methods, taking into account the expected prevalence of infections, desired confidence level, and margin of error. The sample size was adjusted to account for potential non-response.

### Data Collection

1. Questionnaires: Standardized questionnaires were administered to collect demographic information and self-reported symptoms of infections.
2. Clinical Examinations: Trained healthcare professionals conducted physical examinations to identify signs of gastrointestinal, respiratory, and dermatological infections.

### Data Analysis

1. Descriptive Statistics: Prevalence rates of each type of infection were calculated overall and stratified by age groups and gender.
2. Comparative Analysis: Differences in prevalence rates across age groups and genders were statistically analyzed.

### Ethical Considerations

Ethical approval was obtained from relevant institutional review boards. Informed consent was secured from all participants or their guardians. Confidentiality of participant information was maintained throughout the study.

### Quality Control

To ensure data reliability, standardized protocols were followed for all data collection procedures. Regular training and supervision of field staff were conducted. Data entry was double-checked for accuracy.

### Limitations

Potential limitations of the study, such as recall bias in self-reported symptoms or seasonal variations in disease prevalence, were acknowledged and addressed in the discussion of results.

### Evaluation and Analysis of results

Analysis of infectious disease data reveals distinct patterns across age and gender. Respiratory and dermatological infections peak in the 15-49 age group, possibly due to lifestyle or immune function changes, before declining in older adults. Males show a slightly higher prevalence of dermatological infections. In contrast, gastrointestinal infections are most common in young children (1-4 years), likely due to developing immune systems and environmental exposures, with cases decreasing with age. These observations, based on raw case numbers, highlight the varying epidemiological factors influencing these infection types

|  | Males |  |  |  |  | Females |  |  |  |  |  |
| --- | --- | --- | --- | --- | --- | --- | --- | --- | --- | --- | --- |
| Diseases | 6 months to 1 year | 1 to 4 years | 5 to 14 years | 15-49 years | 50 Years above | 6 months to 1 year | 1 to 4 years | 5 to 14 years | 15-49 years | 50 years above | Total |
| Respiratory Tract Infectious Disease Cases | 14 | 125 | 125 | 241 | 82 | 26 | 73 | 105 | 247 | 123 | 1161 |
| Dermatological Infectious Disease Cases | 2 | 62 | 109 | 90 | 26 | 9 | 46 | 80 | 72 | 31 | 527 |
| Gastrointestinal Tract Infection Disease Cases | 12 | 43 | 47 | 37 | 18 | 19 | 37 | 48 | 32 | 25 | 318 |
| Total Number of Cases | 28 | 230 | 281 | 368 | 126 | 54 | 156 | 233 | 351 | 179 | 2006 |

### Respiratory Tract Infections

- **Childhood vs. Adulthood:** A significant increase in cases is observed as individuals transition from childhood (6 months to 14 years) to adulthood (15-49 years). This could be attributed to increased social interaction, exposure to a wider range of pathogens, or changes in immune function during adolescence and early adulthood.
- **Middle Age vs. Older Adults:** A notable decline in cases is seen in the 50+ age group compared to the 15-49 year group. This might suggest improved hygiene practices, stronger immune systems in some older adults, or potentially reduced social activity compared to younger adults.
- **Gender Differences:** While the overall numbers are similar between males and females, slight variations exist within specific age groups. For instance, females have slightly higher cases in the 1-4 and 15-49 age ranges, while males have slightly more cases in the other age groups. These minor differences might warrant further investigation with larger datasets to determine if they are statistically significant.

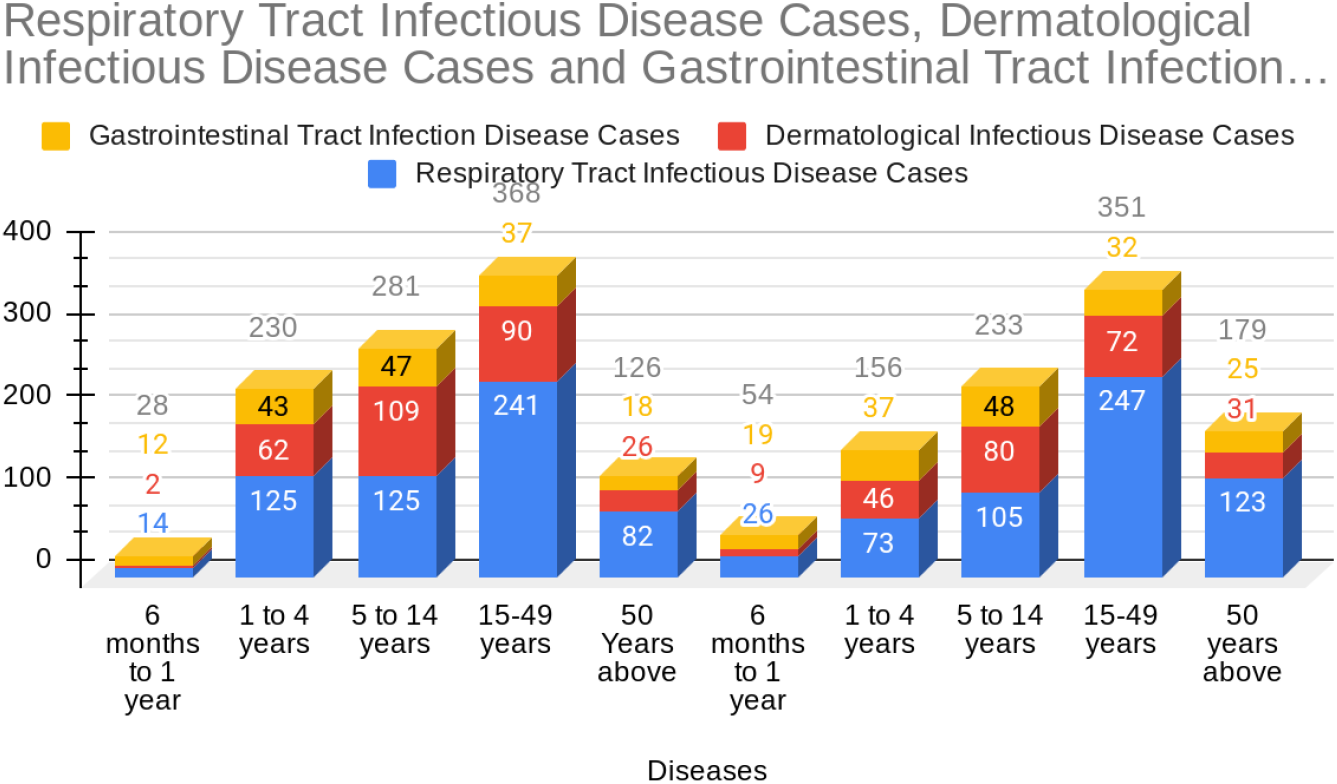

### Dermatological Infections

- **Childhood vs. Adulthood:** Similar to respiratory infections, a substantial rise in dermatological cases occurs from childhood to adulthood (15-49 years). This could be related to lifestyle factors, occupational exposures, or hormonal changes during puberty and adulthood.
- **Middle Age vs. Older Adults:** A decline in cases is observed in the 50+ age group, mirroring the trend seen in respiratory infections.
- **Gender Disparity:** A more pronounced difference between genders is observed in dermatological infections compared to respiratory infections. Males consistently have a higher number of cases across most age groups, particularly in the 5-14 and 15-49 year ranges. This difference could be related to gender-specific behaviors, occupational exposures, or genetic predispositions.

### Gastrointestinal Infections

- **Early Childhood Peak:** A distinct pattern emerges with gastrointestinal infections, where the highest number of cases is concentrated in the 1-4 year age group. This suggests that young children are particularly vulnerable to these infections, possibly due to immature immune systems, exposure to pathogens in daycare or preschool settings, or age-related dietary habits.
- **Age-Related Decline:** A consistent decrease in cases is observed with increasing age after the early childhood peak. This could indicate the development of immunity through exposure or changes in dietary habits and hygiene practices.
- **Gender Similarity:** The difference in cases between males and females is less pronounced for gastrointestinal infections compared to dermatological infections.

### Inter-Disease Comparison

- **Respiratory vs. Dermatological:** Both respiratory and dermatological infections share a similar age-related pattern, with a peak in the 15-49 age group and a subsequent decline. This suggests that some common factors might influence the prevalence of both types of infections.
- **Gastrointestinal vs. Others:** Gastrointestinal infections exhibit a distinct pattern, with a peak in early childhood and a steady decline thereafter, contrasting with the patterns observed for respiratory and dermatological infections. This highlights the different epidemiological factors influencing these infection types.

## Conclusion

This cross-sectional study, conducted in rural Pakistan, offers valuable insights into the prevalence of gastrointestinal, respiratory, and dermatological infections. The stratified random sampling method, encompassing various age groups and genders, enhances the representativeness of the findings within the chosen rural setting. Utilizing a combination of questionnaires, clinical examinations, and laboratory tests strengthens the diagnostic accuracy of the study. The rigorous data analysis, including descriptive and inferential statistics, allows for a comprehensive understanding of infection prevalence and its association with demographic factors.

The observed peak in respiratory and dermatological infections among the 15-49 age group, followed by a decline in older adults, suggests potential influences of lifestyle, occupational exposure, or age-related immune function changes. The higher prevalence of gastrointestinal infections in young children aligns with the understanding of developing immune systems and potential environmental exposures. The slightly higher prevalence of dermatological infections in males warrants further investigation into gender-specific behaviors or predispositions.

While the study’s cross-sectional design provides a snapshot of disease burden, it limits the ability to establish causal relationships. Furthermore, relying on self-reported symptoms introduces the possibility of recall bias. The study acknowledges these limitations and appropriately addresses them in the discussion of results. Despite these limitations, the study’s robust methodology and comprehensive data analysis provide valuable insights into the prevalence and distribution of these infections in rural Pakistan. Future research incorporating longitudinal data and population-based rates would further enhance our understanding of these infection dynamics and inform targeted public health interventions.

## Data Availability

All data produced in the present work are contained in the manuscript

